# Red flags for remote cognitive assessment: An expert consensus study using the Delphi method on behalf of the Canadian Consortium on Neurodegeneration in Aging

**DOI:** 10.1101/2024.10.14.24315468

**Authors:** Nathan H.M. Friedman, Sophie Hallot, Inbal Itzhak, Alex Henri-Bhargava, Jacqueline A. Pettersen, Linda Lee, John D. Fisk, Paula McLaughlin, Vladimir Khanassov, Zahinoor Ismail, Morris Freedman, Howard Chertkow, Richard Camicioli, Philippe Desmarais, Megan E. O’Connell, Maiya R. Geddes

## Abstract

Remote cognitive diagnostic assessment offers numerous benefits, including increased access to care, but it may not always be appropriate, and guidelines are lacking. Our goal was to develop a clinical tool to determine a patient’s suitability for undergoing remote cognitive assessment. A multidisciplinary workgroup, composed of experts in remote assessment, was convened under the auspices of the Canadian Consortium on Neurodegeneration in Aging. Delphi, an anonymous group consensus method, was used to determine ‘red flags’ for remote cognitive diagnostic assessment. The process consisted of one round of flag generation, then two rounds of flag scoring based on effectiveness, reproducibility, and efficiency. In the first round, 11 respondents generated 67 potential flags. In subsequent rounds, 8 and 9 respondents, respectively, scored the flags, yielding 14 red flags that met the predetermined consensus criteria. This research led to the creation of a novel clinical decision-making infographic to support multidisciplinary clinicians in determining a patient’s readiness to undergo remote cognitive and behavioral diagnostic assessment.

**Highlights:**

- A timely and accessible diagnosis of dementia is crucial for optimal patient care.
- Although clinicians are increasingly using telemedicine, guidelines on patient suitability for remote cognitive and behavioral assessment are lacking.
- To address this knowledge gap, we developed a clinical decision-making tool to determine if a cognitive evaluation via telemedicine should be avoided.
- To synthesize expert opinion, we used the Delphi method, an anonymous group consensus method that reduces eminence bias.
- In collaboration with knowledge translation experts, an infographic describing the final 14 red flags for remote cognitive assessment was developed for clinicians to determine the appropriateness of patients for remote dementia diagnostic assessment.

**Research in context:** *Systematic Review:* PubMed and Google Scholar were used to survey the literature. Our review found that although there was a need for better access to dementia care, a framework to provide such care remotely is in development. We identified a gap in clinical guidelines on contraindications for remote cognitive assessment.

*Interpretation:* This study created a clinical decision-making tool as an accessible infographic for clinicians to use when considering a remote cognitive diagnostic assessment. This guideline is based on expert consensus.

*Future Directions:* In future studies, patient and caregiver perspectives should be incorporated into the decision-making process, and this tool should be validated in clinical contexts. Some of the identified flags for remote assessment are modifiable, and strategies to mitigate burden on patients and caregivers are warranted.

## Background

The number of adults aged 60 or over is expected to double by 2050 to 2.1 billion people worldwide [1]. Dementia is critically linked to aging and currently affects an estimated 55 million people globally; this is expected to affect 78 million people by 2030 [2]. A timely diagnosis of dementia is essential for patient care, empowerment, future planning, and access to services [3]. Furthermore, emerging therapies are dependent on correctly identifying patients at early stages of neurodegenerative disease. The diagnostic approach to dementia begins with a neurobehavioral status exam to establish a diagnosis [4,5]. A comprehensive cognitive and behavioral evaluation leading to a diagnosis of dementia involves an encounter between a healthcare professional and the patient, with essential collateral input from the caregiver [6]. After a diagnostic impression is formed, a care plan can then be developed which may include additional investigations to establish the diagnosis and characterize the risk of clinical progression [7].

Telemedicine is defined by the Centers for Medicare & Medicaid Services as the “two-way, real time interactive communication between the patient and the physician or practitioner at [a] distant site” [8]. Telemedicine has long been used to assess patients, such as those living in remote areas [9–11]. Prior research has shown that adults in rural communities may be underdiagnosed or diagnosed later than their urban counterparts [12]. The recent Coronavirus Disease 2019 (COVID 19) pandemic led to a dramatic rise in the use of telemedicine because of social distancing guidelines. A virtual dementia care framework was created by the Alzheimer Society of Canada Task Force on dementia care best practices for COVID-19 and beyond [13]. In the past few years, we have seen rapid development of novel tools and guidelines aimed at integrating virtual care into the existing healthcare infrastructure [13–16] and more widespread acceptance of telemedicine in medical practice more generally [17]. Telemedicine used for remote cognitive assessment has been proposed as an accessible approach that can be scaled to population-level screening, in combination with blood-based biomarkers [18]. However, remote neurobehavioral assessment has important limitations that should be carefully considered in its implementation, including potential negative impacts on the therapeutic alliance between patient and physician, technical hurdles, and the potential to widen existing healthcare disparities [19]. These underline the importance of identifying the appropriate context for remote cognitive assessment, and situations when remote cognitive assessment should be avoided.

To fulfill competency of care in telemedicine, clinician proficiency with technology and awareness of its limitations are required [20,21]. For example, one cross-sectional study found that around 60% of neurologists required technical support while engaging in telemedicine [22]. Systematic reviews of remotely administered cognitive assessments have found that patient factors including older age, slower internet connection speed, scoring close to cutoff on screening tests, and tasks requiring a greater motor response (e.g., clock drawing) limited the validity of the remotely delivered test results [23,24]. Critically, however, there is currently no consensus on which other factors may render a remote Neurobehavioral Status Exam inappropriate and warrant a switch to in-person assessment [21,23,25–27].

Our goal was to establish a guidance framework to support multidisciplinary physicians in identifying situations when it may be best not to perform a cognitive and behavioral diagnostic assessment remotely. To achieve this goal, we developed a clinical decision-making tool to identify features about the patient, caregiver, clinician, or context that would suggest the need to shift to an in-person encounter. The target audience for this tool is multidisciplinary physicians who engage in remote diagnostic assessment of cognitively impaired adults. To develop this tool and infographic, the Delphi method was employed to synthesize multidisciplinary expert opinion from a workgroup convened under the auspices of the Canadian Consortium on Neurodegeneration in Aging (CCNA) [28–30]. The Delphi method is an iterative process to systematically establish group consensus while mitigating potential bias [28]. Next, we engaged with knowledge translation experts to develop an infographic in English and French. By creating a clinical decision-making tool that is evidence-based and openly accessible, we hope to meaningfully impact the clinical care and diagnostic validity of people living with cognitive impairment.

## Methods

### Workgroup creation

The CCNA telemedicine workgroup came together under the auspices of the CCNA and was tasked with advancing guidance for remote cognitive assessment and diagnosis in dementia care. In forming the group, steps were taken to ensure multidisciplinary participation by inviting experts throughout the CCNA network. The workgroup within the CCNA was composed of an expert multidisciplinary team of behavioral neurologists, neuropsychiatrists, neuropsychologists, social workers, geriatricians, and family physicians.

### Study design

In our study, the Delphi method was used to find consensus amongst the telemedicine workgroup within the CCNA. This method is particularly useful in the medical field, where evidence is often conflicting or insufficient [28,29]. The main advantages of the method are that it avoids eminence bias, can be performed remotely and asynchronously at low cost, and allows individuals to consider the group opinion when formulating a response. In a three-round process, our participants first proposed data, then evaluated and re-evaluated data (Figure 1).

**Figure 1 –.**
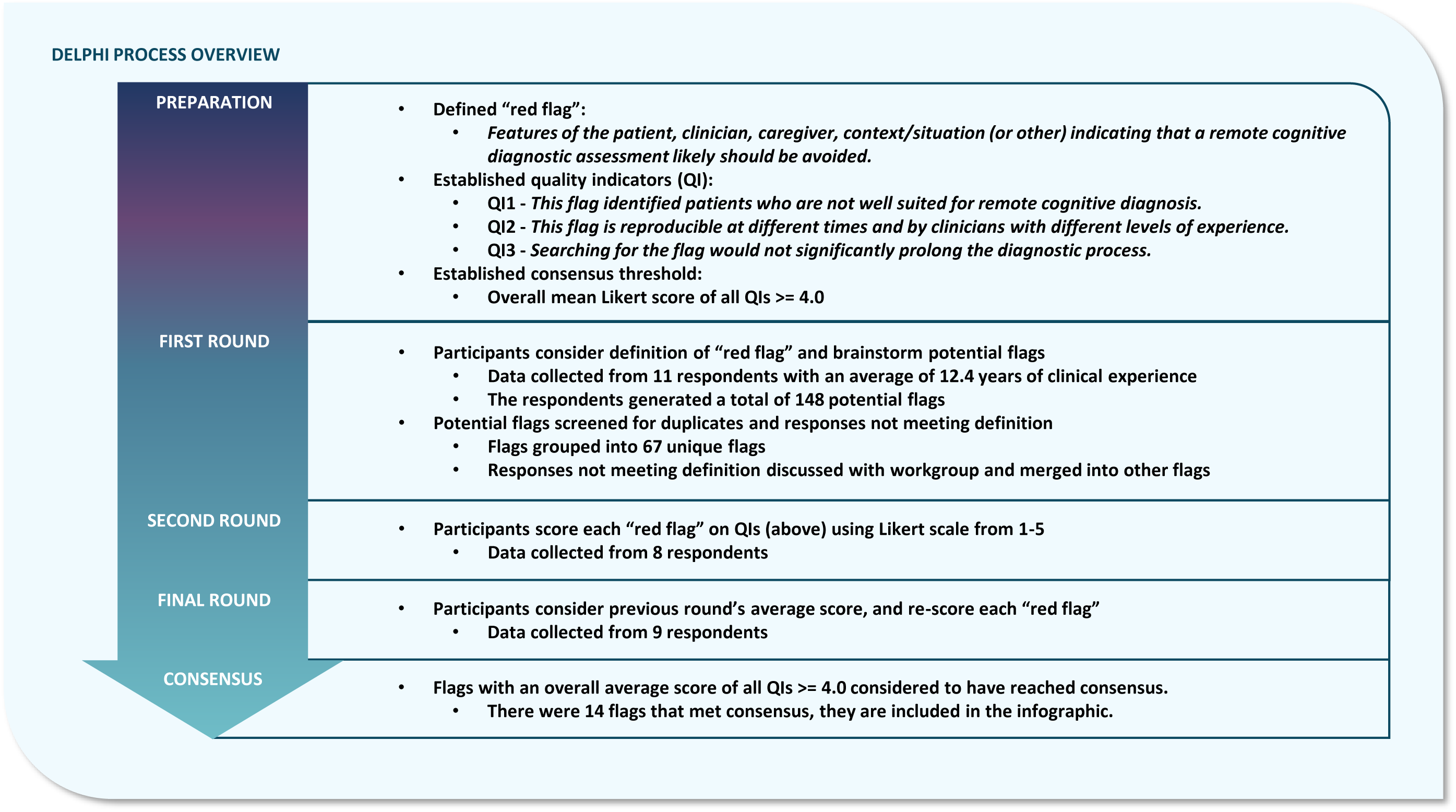
Overview of the Delphi process implemented in the current study.

The study took place between January 19, 2023, and September 20, 2023. Data collection for each round was performed using web-based surveys (Microsoft Forms). Though the surveys were distributed to a closed group, responses were anonymous, ensuring each response had equal weight.

### Establishing definitions, process, and consensus criteria

An initial virtual meeting with the workgroup served to establish the necessary prerequisites for the Delphi process [28]: This included the definition of ‘red flag’, the quality indicators for scoring candidate flags, and the consensus threshold. The goal was to have a definition of ‘red flag’ that was simple, comprehensive and encompassed the important components of a clinical encounter. The agreed upon definition of a ‘red flag’ was “Features of the patient, clinician, caregiver, context/situation (or other) indicating that a remote cognitive diagnostic assessment likely should be avoided.”

Three quality indicators (QIs) were used to score each flag quantitatively on a 5-point Likert scale [31]. The goal in choosing the indicators was to capture the important attributes of a red flag in as few indicators as possible with clear language. The first QI focused on efficacy, “This flag identified patients who are not well suited for remote cognitive diagnosis.” The second QI focused on reproducibility, “This flag is reproducible at different times and by clinicians with different levels of experience.” The third QI examined feasibility, “Searching for this flag would not significantly prolong the diagnostic process.” Each potential response on the Likert scale was rated as follows: strongly disagree=1, disagree=2, neutral=3, agree=4, strongly agree=5.

Consensus criteria were set a priori to reduce the risk of bias. We considered consensus to be met when the average score for all QIs taken together for a specific potential red flag was 4.0 or greater.

### The Delphi process

The first round of the Delphi generated all the potential flags to be voted on in later rounds. It was important to capture a set of flags that covered the spectrum of opinions held by the group, as later steps generally serve to filter out flags that did not meet consensus. We designed the prompts to be as inclusive as possible and stimulate brainstorming. We prompted respondents about important components of a clinical encounter including features of the patient, caregiver, patient, or context (e.g., “What are red flags about the patient’s [caregiver] that indicate a remote assessment should be avoided?”). Participants responded anonymously by generating as many potential red flags as possible. Basic demographic information about the expert participants, including their specialty and years of clinical experience was also collected in this round.

After the first round, all the responses were pooled and screened for validity (whether the potential factor met the agreed up upon definition of “red flag”) and duplicate responses. With CCNA telemedicine workgroup feedback and approval, invalid responses were removed, and duplicate responses were combined under the response with the clearest language. The workgroup concluded that all remaining potential red flags in the set were valid.

In the second round, respondents were shown each potential red flag and asked to score it based upon the three QIs using the above Likert scale. At the end of the round, respondents were given the opportunity to recommend new red flags that they felt were missing at this point, and one additional flag was generated overall. After the second round, the group average scores for each red flag were calculated, including the average for each QI and overall.

In the third round, respondents were shown each red flag again along with the group average scores from the previous round, for each QI and overall, for all but the one flag added in the second round. They were asked to re-score each flag using the same QI Likert scale described above while considering the previous round’s averages.

After the third round, the group average scores for each flag were re-calculated. The consensus threshold was applied (average score for all QIs≥4.0). Flags with scores above the consensus threshold were included in the final set of agreed upon red flags.

### Infographic

In collaboration with a knowledge translation specialist from the CCNA Operations Center, the final set of red flags was translated into a user-friendly infographic as a clinical decision-making tool. This tool was designed to aid physicians in determining if a patient is an appropriate candidate for cognitive diagnostic assessment via telemedicine. If one of the final red flags is identified before a clinical encounter, then the patient is likely not well suited for cognitive assessment via telemedicine, and switching to in-person assessment is recommended.

## Results

### Statistical analysis

In the first round of the Delphi process, there were 12 respondents, possessing an average of 14.6±11.1 years (mean ± standard deviation) of clinical experience within neurology, psychiatry, psychology, social work, geriatrics, and health administration. This group generated a total of 151 flags. Only 3 were considered invalid based on the definition criterion. Of the 148 remaining red flags, 82 were duplicated flags that coalesced into the 66 unique candidate red flags.

In the second round, 8 respondents scored each candidate red flag on the three QIs (Supplemental Table 1). One additional flag was added in this round, which was a modification of a prior candidate red flag, “The patient does not have access to a caregiver during the remote assessment.” The new candidate flag from the second round was “The patient does not have access to a caregiver during the remote assessment AND is more than mildly impaired.”

In the third round, 9 respondents re-scored each flag while considering the previous round’s scores. The mean score and standard deviation for each potential red flag are shown in Table 1. Applying the consensus criterion after this third and final round yielded 14 red flags that scored over threshold, shown in bold at the top of Table 1. An infographic of the synthesized red flags was developed in collaboration with a knowledge translation specialist within the CCNA Operations Center to provide decision-making guidance about the suitability of a patient for remote cognitive assessment (Figure 2). If any of the 14 red flags in the decision-making tool are identified, a decision to shift to an in-person clinical assessment is warranted.

**Figure 2 –.**
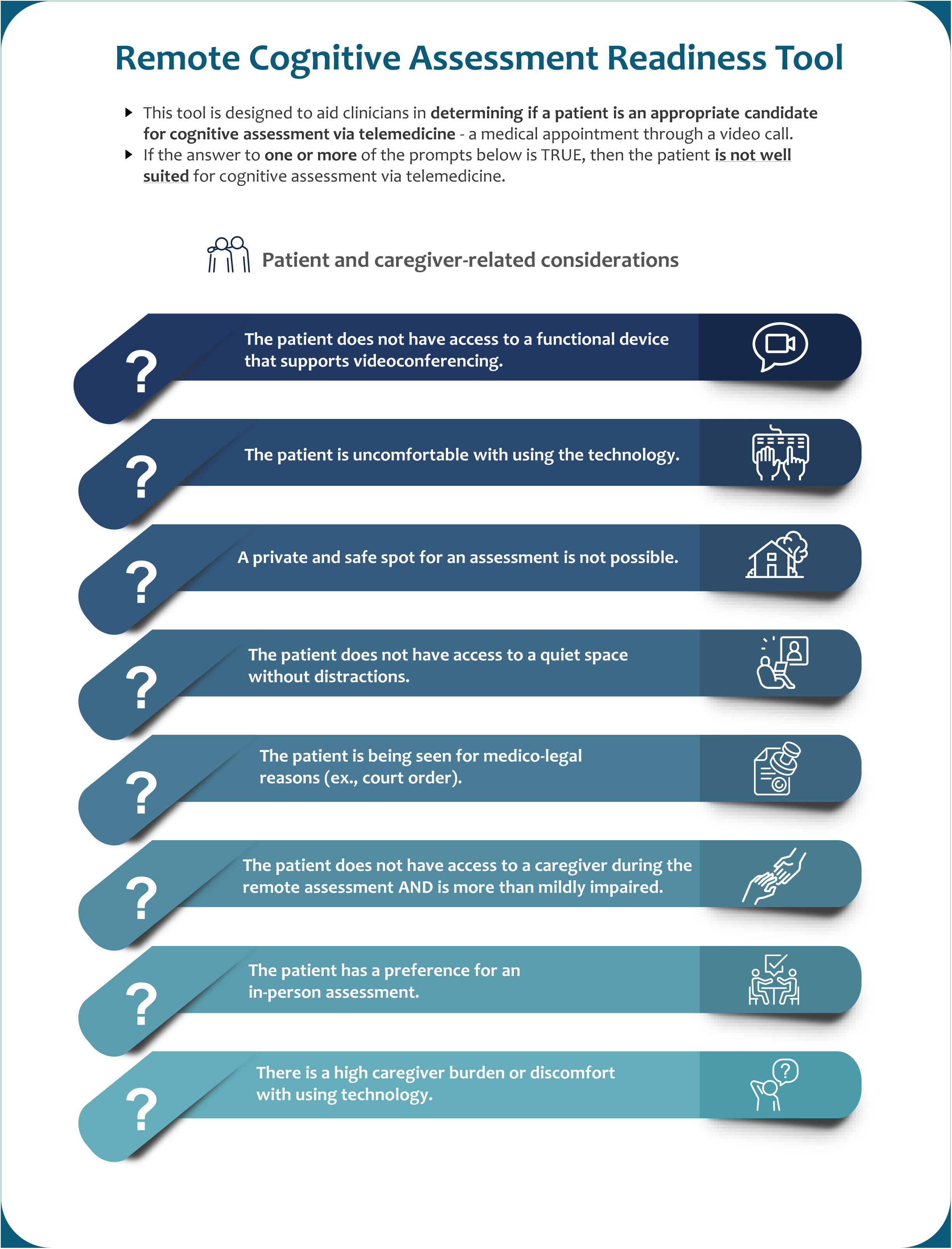
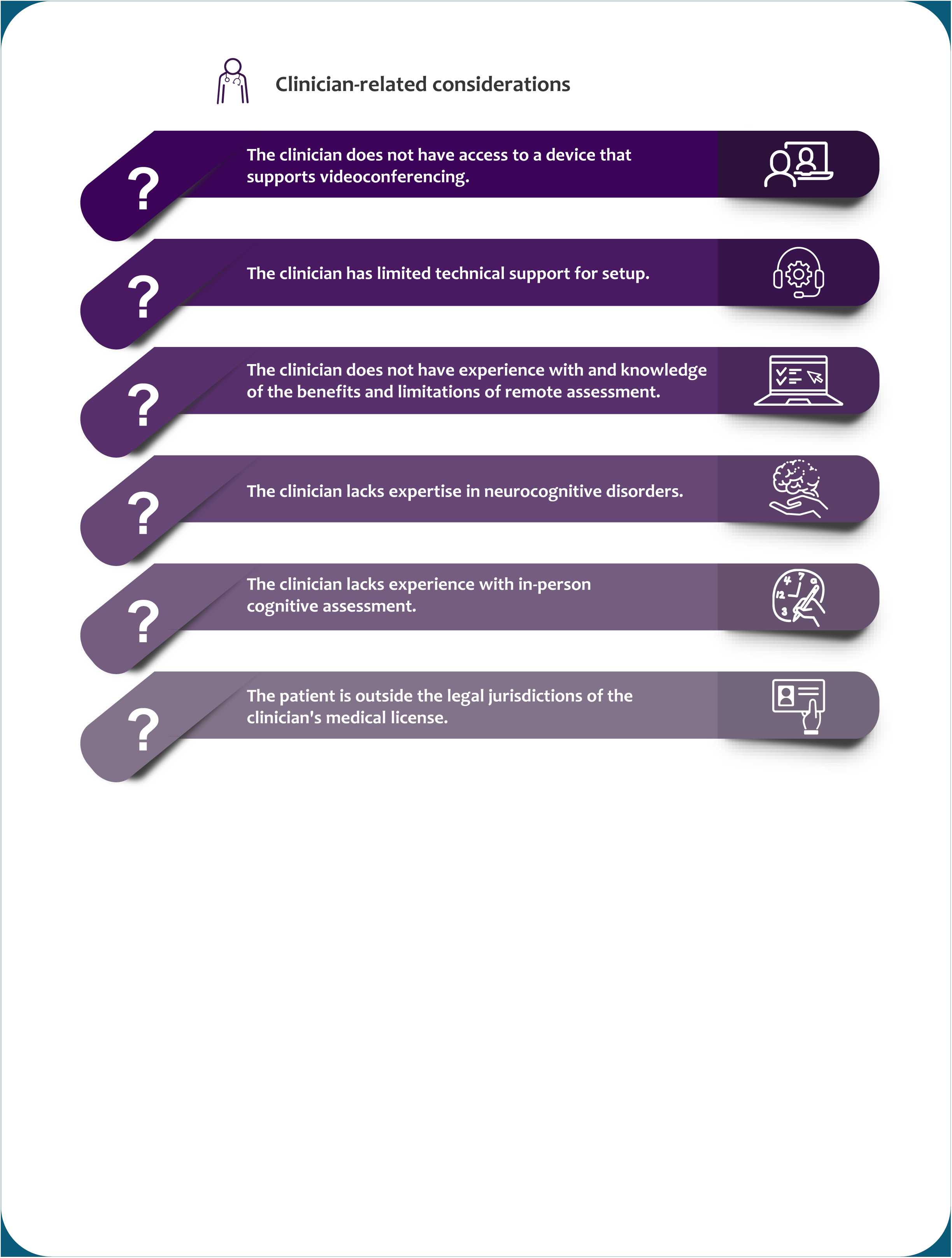
The infographic and clinical decision-making tool.

**Table 1 –.** List of all the red flags with quality indicator (QI) statistics from the respondents after the third and final round of the Delphi process. In this table, the flags are sorted in descending order of mean score. The flag number in the leftmost column represents the order in which the flags were shown to respondents. The statistics shown are mean +/- standard deviation (SD). The first 14 flags on this table (bolded) reached consensus as the overall mean was >=4.0.

| Red Flag |  | QI1 | QI2 | QI3 | Overall |
| --- | --- | --- | --- | --- | --- |
| | | Mean±SD | Mean±SD | Mean±SD | Mean±SD |
| **1** | **The clinician does not have access to a device that supports videoconferencing.** | **4.9±0.3** | **4.8±0.4** | **5.0±0.0** | **4.9±0.3** |
| **2** | **The patient is outside the legal jurisdictions of the clinician's medical license.** | **5.0±0.0** | **4.8±0.4** | **4.9±0.3** | **4.9±0.3** |
| **3** | **The patient does not have access to a functional device that supports videoconferencing.** | **4.9±0.3** | **5.0±0.0** | **4.3±0.7** | **4.7±0.4** |
| **4** | **A private and safe spot for an assessment is not possible.** | **4.9±0.3** | **4.4±0.5** | **4.3±0.7** | **4.6±0.5** |
| **5** | **The clinician lacks expertise in neurocognitive disorders.** | **4.7±0.5** | **4.3±0.5** | **4.4±0.5** | **4.5±0.5** |
| **6** | **The patient does not have access to a quiet space without distractions.** | **4.8±0.4** | **4.4±0.5** | **4.2±0.4** | **4.5±0.4** |
| **7** | **The clinician has limited technical support for setup.** | **4.4±0.5** | **4.3±0.5** | **4.4±0.5** | **4.4±0.5** |
| **8** | **The patient is being seen for medico-legal reasons (ex., court order)** | **4.0±0.9** | **4.3±0.5** | **4.3±0.5** | **4.2±0.7** |
| **9** | **The clinician does not have experience with and knowledge of the benefits and limitations of remote** | **4.3±0.7** | **3.9±0.6** | **4.2±0.6** | **4.1±0.6** |
| **10** | **The patient has preference for in-person assessment.** | **4.0±0.7** | **4.1±0.6** | **4.0±0.9** | **4.0±0.7** |
| **11** | **There is high caregiver burden or discomfort with using technology.** | **4.2±0.6** | **4.0±0.5** | **3.9±0.7** | **4.0±0.6** |
| **12** | **The patient does not have access to a caregiver during the remote assessment AND is more than mildly** | **4.6±0.5** | **3.9±0.9** | **3.7±1.1** | **4.0±0.8** |
| **13** | **The clinician lacks experience with in-person cognitive assessment.** | **4.0±0.9** | **4.1±0.7** | **4.0±0.7** | **4.0±0.8** |
| **14** | **The patient is uncomfortable with using the technology.** | **4.1±0.6** | **4.1±0.6** | **3.8±0.8** | **4.0±0.6** |
| 15 | The clinician does not have access to the patient's prior chart or imaging. | 3.6±0.8 | 4.0±0.8 | 4.2±0.4 | 3.9±0.7 |
| 16 | There are language barriers or the standardized assessments are not available in the patient's native | 3.8±0.6 | 4.1±0.3 | 3.9±0.7 | 3.9±0.6 |
| 17 | The patient has moderate-severe sensory impairment (ex., hearing loss). | 4.0±0.7 | 4.0±0.5 | 3.7±0.7 | 3.9±0.6 |
| 18 | The patient is suspected to have delirium or altered level of consciousness. | 4.7±0.5 | 3.7±0.7 | 3.2±0.8 | 3.9±0.7 |
| 19 | The caregiver has preference for in-person assessment. | 4.0±0.7 | 3.8±0.4 | 3.8±0.8 | 3.9±0.6 |
| 20 | The clinician is distracted or lacks time to properly perform the assessment. | 4.1±0.9 | 3.4±1.1 | 3.9±0.9 | 3.8±0.9 |
| 21 | The patient does not have access to a caregiver during the remote assessment. | 3.2±0.9 | 4.1±0.6 | 4.0±0.8 | 3.8±0.8 |
| 22 | The patient or caregiver will not have technical support at the time of the encounter. | 3.6±0.8 | 3.8±0.8 | 3.9±0.7 | 3.7±0.8 |
| 23 | The patient has an atypical presentation that requires more thorough neuropsychological testing. | 3.8±0.9 | 3.6±0.5 | 3.8±0.9 | 3.7±0.8 |
| 24 | It is the first diagnostic assessment to address cognitive or behavioural concerns. | 2.2±0.9 | 4.3±0.5 | 4.2±0.4 | 3.6±0.6 |
| 25 | The patient is being seen for a second opinion. | 2.1±0.7 | 4.2±0.4 | 4.2±0.4 | 3.5±0.5 |
| 26 | The caregiver is unable to participate in a separate private interview. | 3.3±0.5 | 3.6±0.8 | 3.7±0.8 | 3.5±0.7 |
| 27 | There is suspicion of abuse (physical, psychological, sexual, financial) or neglect from the caregiver. | 4.7±0.5 | 3.3±0.5 | 2.4±0.5 | 3.5±0.5 |
| 28 | The patient has significant anxiety from the computer screen or seeing themselves on the screen. | 3.8±0.9 | 3.2±0.6 | 3.2±0.8 | 3.4±0.8 |
| 29 | The assessment is at a time of day when the patient might not be awake or alert. | 3.2±0.8 | 3.9±0.6 | 3.1±1.0 | 3.4±0.8 |
| 30 | It would be impossible to eventually examine the patient in-person. | 2.6±1.0 | 4.0±0.8 | 3.7±0.9 | 3.4±0.9 |
| 31 | The clinician does not have access to a service corridor for the patient. | 3.3±0.8 | 3.4±1.0 | 3.3±1.1 | 3.4±0.9 |
| 32 | The caregiver is opposed to cognitive assessment of the patient. | 3.4±0.8 | 3.4±0.7 | 3.1±0.9 | 3.3±0.8 |
| 33 | The clinician lacks the skills to diffuse unexpected behavior from the patient or caregiver. | 3.8±0.6 | 3.1±0.9 | 3.1±0.9 | 3.3±0.8 |
| 34 | The patient is easily distracted, fidgeting, or walking away from the computer. | 3.3±0.5 | 2.9±0.7 | 3.7±0.8 | 3.3±0.7 |
| 35 | The clinician lacks patience and empathy. | 3.7±0.8 | 3.1±1.0 | 3.1±1.0 | 3.3±0.9 |
| 36 | The patient scores close to cutoff on a standardized cognitive assessment. | 2.4±1.0 | 3.9±0.7 | 3.6±0.8 | 3.3±0.8 |
| 37 | The patient does not have sufficient ability to move around as needed (ex., the patient is bedbound.) | 2.0±0.9 | 4.0±0.8 | 3.8±0.8 | 3.3±0.9 |
| 38 | Disclosure of the diagnosis or consequences thereof is needed during the encounter. | 2.4±0.8 | 3.8±0.8 | 3.4±1.0 | 3.2±0.9 |
| 39 | The remote cognitive assessment is not in the best interest of the patient. | 4.2±0.9 | 2.8±0.6 | 2.7±0.8 | 3.2±0.8 |
| 40 | The patient is suspected to have a rapidly progressive neurodegenerative disease. | 2.8±1.2 | 3.4±0.5 | 3.3±0.7 | 3.2±0.9 |
| 41 | The patient has significant language dysfunction (ex., primary progressive aphasia) | 2.7±1.2 | 3.6±0.8 | 3.2±1.1 | 3.1±1.1 |
| 42 | The patient has a known medical or psychiatric condition that is decompensated. | 3.6±0.7 | 3.1±0.7 | 2.7±1.1 | 3.1±0.8 |
| 43 | The caregiver is known to be unreliable. | 3.4±0.7 | 2.9±0.6 | 2.7±0.9 | 3.0±0.7 |
| 44 | The patient's environment contains orienting cues. | 3.2±0.6 | 3.1±0.7 | 2.7±0.8 | 3.0±0.7 |
| 45 | The patient does not have a long term residence or moves residences frequently. | 2.6±1.0 | 3.3±0.8 | 3.0±0.7 | 3.0±0.8 |
| 46 | The patient has developed racial bias, and is abusive towards the clinician. | 3.0±0.5 | 3.1±0.6 | 2.8±0.6 | 3.0±0.6 |
| 47 | The patient has severe visuospatial deficits. | 3.4±0.8 | 3.1±0.6 | 2.2±0.6 | 2.9±0.7 |
| 48 | The patient has significant uncontrolled pain. | 2.3±0.9 | 3.2±0.6 | 3.2±0.9 | 2.9±0.8 |
| 49 | There are physical manifestations accompanying the cognitive impairment, necessitating an in-person | 3.0±0.9 | 2.9±0.7 | 2.9±0.7 | 2.9±0.8 |
| 50 | The caregiver is a temporary caregiver. | 2.3±0.7 | 3.0±0.7 | 3.4±0.5 | 2.9±0.6 |
| 51 | The caregiver is uncomfortable discussing matters in front of the patient. | 3.0±0.7 | 2.9±0.9 | 2.9±0.9 | 2.9±0.8 |
| 52 | The patient has severe behavioural issues. | 2.2±0.9 | 3.4±0.7 | 3.0±0.7 | 2.9±0.8 |
| 53 | The patient is too cognitively impaired to understand the process of video assessment. | 3.4±1.1 | 2.9±0.7 | 2.3±0.9 | 2.9±0.9 |
| 54 | The patient has mild-moderate, or worse, psychotic symptoms. | 2.1±0.7 | 3.4±0.7 | 3.1±0.6 | 2.9±0.7 |
| 55 | The caregiver is known to have unstable medical or psychiatric disease. | 3.2±0.4 | 2.8±0.6 | 2.7±0.8 | 2.9±0.6 |
| 56 | The caregiver is distrustful of the clinician. | 3.4±0.7 | 2.7±0.7 | 2.6±0.7 | 2.9±0.7 |
| 57 | There has been a sudden change in the patient's clinical status. | 2.9±0.6 | 2.7±0.5 | 3.0±0.5 | 2.9±0.5 |
| 58 | The patient consumes alcohol regularly or at unusual times of the day. | 2.3±1.2 | 3.2±0.8 | 2.8±0.6 | 2.8±0.9 |
| 59 | The patient has suicidal ideas. | 2.9±1.2 | 3.0±0.7 | 2.3±0.8 | 2.7±0.9 |
| 60 | The caregiver is interrupting the interview frequently. | 3.1±0.7 | 2.9±0.7 | 2.2±0.8 | 2.7±0.8 |
| 61 | The clinician feels they must censor themselves. | 3.1±0.7 | 2.7±0.7 | 2.4±0.5 | 2.7±0.6 |
| 62 | The caregiver is suspected to have cognitive impairment. | 3.0±1.2 | 2.7±0.8 | 2.3±0.7 | 2.7±0.9 |
| 63 | The patient has severe anosognosia. | 2.6±0.8 | 2.8±0.6 | 2.3±0.5 | 2.6±0.7 |
| 64 | The caregiver is showing signs of significant burnout. | 2.1±0.9 | 2.9±0.7 | 2.6±0.8 | 2.5±0.8 |
| 65 | The caregiver has racial bias towards the clinician. | 2.2±0.8 | 2.3±0.8 | 2.6±0.8 | 2.4±0.8 |
| 66 | The caregiver has lots of concerns that need to be addressed. | 1.8±0.6 | 2.9±0.9 | 2.3±0.7 | 2.3±0.7 |
| 67 | The caregiver is anxious. | 1.8±0.4 | 2.4±0.8 | 2.2±0.6 | 2.1±0.6 |

### Infographic

The goal of this study was to create a clinical decision-making tool to determine a patient’s readiness for remote cognitive assessment (Figure 2). If a health professional identifies any of the 14 red flags, a shift to an in-person assessment is preferred. The tool was developed in English and translated to French.

## Discussion

In this study, the Delphi method established consensus among experts regarding patient suitability for remote cognitive and behavioral assessment. This approach resulted in fourteen red flags that reached consensus. These flags were features of the patient, caregiver, clinician or context of the clinical encounter that suggest a shift to in-person assessment should be considered. The flags were translated into a user-friendly infographic and clinical decision- making tool accessible by clinicians considering the suitability of a given patient for a diagnostic neurobehavioral assessment via telemedicine.

### The Delphi method

The Delphi method is versatile and has been used in various domains of research including finance, economics, process planning, and healthcare [28,29]. Most examples of the method retain the core elements of anonymity, iteration, feedback, and consensus, with validity of the results depending on the implementation. [28,29].

In this study, the overall design and consensus criteria were defined prior to beginning the Delphi iterative rounds. Like most studies in healthcare [29], our study used a predetermined, rather than variable, number of rounds. Our aim was to achieve a balance between allowing participants to respond to the group and reducing response fatigue. Setting a predetermined consensus threshold is recommended as it increases transparency within the group, since participants are aware how close a flag is to achieving consensus [29]. This approach also reduces the subjectivity of study operators.

The iterations in this study were carried out using online software, which allowed us to gather de-identified data and uphold the principle of anonymity. This is important to minimize the risk of individuals having disproportionate influence on the group, known as eminence bias. The online forms also allowed data collection in the context of a national workgroup where individuals were dispersed across the country, and working asynchronously over a wide range of time. Our aim in having asynchronous rounds was to minimize the burden on respondents and maximize response rate.

### The red flags

The majority of the red flags that reached consensus acknowledge the importance of patient and caregiver factors that could make a remote assessment inappropriate (Flags 2, 3, 4, 6, 8, 10, 11, 12, 14, Table 1). This includes flags focused on comfort and familiarity with technology and access to electronic devices and internet connectivity (Flags 3, 11, 14, Table 1). The importance of logistical and technical considerations has been identified as a limiting factor for people living in rural or remote areas, as those without access to high-speed internet had reduced access to telemedicine during the COVID-19 pandemic [32]. In the context of remote cognitive assessment, a cross-sectional study cited limited access and technological understanding as barriers to participation [33]. Most studies evaluating remote cognitive assessment have used remote medical hubs with an existing technology infrastructure, and this is reflected in new models of telemedicine for remote cognitive assessment [26,34]. There are also many programs to increase access to high-speed internet, such as *Internet for All* in the United States, and the *Universal Broadband Fund* in Canada [35,36]. This highlights the potentially modifiable nature of certain red flags, whereby changes in community access and healthcare policy could potentially reduce the barriers to remote cognitive assessment.

The use of accessible satellite clinics could potentially address red flags identified in the current study, including access to a private, safe location, and a location that is quiet and free of distractions (Flags 4, 6, Table 1). Privacy during a medical encounter is critical for patient autonomy, where a patient is free to decide who is present in a clinical encounter. Additionally, the presence of distractions impedes cognitive performance [37], thereby reducing testing validity. Patient preference is significant; if a patient prefers an in-person assessment, this should be respected (Flag 10, Table 1).

A systematic review of remote cognitive assessment identified the presence of a caregiver as a critical facilitating factor [39]. Multiple flags identified by the workgroup underscore the critical role played by caregivers (Flags 11, 12, Table 1). One study comparing in-person and remote cognitive assessment in a memory clinic found that 68% of patients required caregiver assistance to participate in a videoconference clinical encounter [40]. Another study showed that patients with reduced cognitive function required greater caregiver involvement to complete remotely administered computerized and standard cognitive testing [41].

Current legal guidelines allow for remote cognitive assessment, but practicing outside of the jurisdiction of a clinician’s license has implications for patient privacy, quality of care, clinician liability, and remuneration [38]. Thus, legal considerations are essential to determine the appropriateness of a remote diagnostic encounter (Flag 2, Table 1). Furthermore, the group identified that a court-ordered assessment is sufficient reason to switch to an in-person assessment (Flag 8, Table 1). An in-person neurobehavioral assessment ensures that the clinician is less reliant on factors and people in the patient’s environment that may erroneously influence the diagnostic impression.

One flag that came close to, but did not reach, consensus was the presence of sensory impairment (Flag 17, Table 1). Efficacy and inter-rater reliability scored highly; however, potential challenges in collecting this information efficiently prevented this flag from achieving consensus. Previous studies have shown that participants with vision or hearing impairment have lower performance on cognitive assessments, even when adjusting for the real or simulated impairment [42–45]. Sensory impairments are increasingly common with age [1], and the presence of sensory impairment was often an exclusion criterion for studies focused on remote cognitive assessment ￼[46]￼. Standardized remote methods of screening for visual impairment have been established, yet the accuracy and reliability of hearing impairment remote screening methods are not ￼￼[47,48]￼. This suggests that valid strategies for rapid, remote screening of hearing and vision represent a unmet clinical need that would help to optimize patient suitability for remote cognitive assessment.

The group identified several clinician-related flags (Flags 1, 5, 7, 9, 13, Table 1). A fundamental prerequisite for remote cognitive assessment is the clinician’s access to secure, confidential software and appropriate technology (Flags 1, 7, Table 1) [49]. However, this can be costly and often requires administrative support that poses a potential barrier [50]. The group also highlighted the importance of clinician expertise to the validity of the remote clinical encounter (Flags 5, 9, 13, Table 1). Competency in performing remote cognitive assessment includes both knowledge of the benefits and limitations of telemedicine and expertise in neurocognitive disorders in an in-person context [13,20]. A systematic review on barriers to dementia care among primary care physicians identified concerns about knowledge of dementia, lack of training in dementia care, and discomfort with conducting cognitive assessments [51]. While training clinicians in neurobehavioral assessment is anchored by in-person learning, remote assessment is also a skill requiring exposure and practice. Early examples have shown that including telemedicine training in residency programs is desired [52], and that optimizing the residency curriculum to include training in remote care benefited residents’ competency and patient access to care [53,54].

### Limitations and future directions

While this research provides guidance on virtual cognitive assessment readiness, it is important to be aware of some of the limitations of this work. First, our findings are based on expert consensus and this guidance and clinical decision-making tool have not yet been validated in real-world clinical settings. Future work could integrate feedback from clinicians that would allow for estimation of the sensitivity and specificity of each flag, along with quantitative measures of reproducibility and feasibility. Second, as with any clinical tool, information from these guidelines must be interpreted within the clinical context. This tool does not replace clinical judgment, and the clinician must make the final decision whether telemedicine is appropriate (Flag 9, Table 1).

To enhance the validity of this novel clinical decision-making tool, future studies could aim to incorporate a wider range of stakeholder perspectives. The perspectives of patients, caregivers, and other persons with lived experience on what factors warrant an in-person assessment are fundamental to providing patient-centered care.

Some of the identified red flags are potentially modifiable and represent opportunities for improving healthcare guidelines and advocacy efforts. Utilizing remote cognitive assessment for referral and diagnosis appropriately has the potential to increase access via population screening programs, as proposed by a recent study in Sweden [18]. Easily accessible internet and technological support could reduce caregiver and patient burden. Rural access to high-speed internet and health resource centers offering internet-enabled, quiet, and secure spaces are proven methods for achieving this goal [34,46,55]. The cost-benefit analysis in creating such centers should be considered against the healthcare system advantages that telemedicine affords [56]. These efforts have important public health implications with an aging population that is living longer, and work has already begun by governments to reduce the digital divide [35,36].

## Conclusion

The Delphi method was used to synthesize expert opinion regarding patient suitability for remote cognitive diagnostic assessment into a clinical decision-making tool consisting of red flags about the patient, caregiver, clinician, or context that potentially render a remote assessment invalid. This tool addresses the need for evidence-based decision-making guidance about a patient’s suitability for remote assessment as an infographic to maximize knowledge translation into clinical practice. With the increasing use of telemedicine in the context of cognitive and behavioral health, a user-friendly and openly accessible tool will help patients receive appropriate care and help clinicians avoid potential pitfalls associated with remote assessment. We hope that this work will catalyze public health advocacy and research to better understand how barriers to telemedicine can be further mitigated.

## Data Availability

All data produced in the present study are available upon reasonable request to the authors.

## Acknowledgements and funding

We are grateful for the administrative support of Shoshana Green, Annie Le Bire, Maria Di Nezza and to the Canadian Consortium on Neurodegeneration in Aging.

This research was made possible by funding from a National Sciences and Engineering Research Council of Canada (NSERC) Discovery Grant (DGECR-2022-00299), an NSERC Early Career Researcher Supplement (RGPIN-2022-04496), a Fonds de Recherche Santé Québec (FRSQ) Salary Award, the Canada Brain Research Fund (CBRF), an innovative arrangement between the Government of Canada (through Health Canada) and Brain Canada Foundation, the Alzheimer Society Research Program (ASRP) New Investigator Grant, the Brain Canada Future Leaders Award, the Canada First Research Excellence Fund, awarded through the Healthy Brains, Healthy Lives initiative at McGill University, and the Canadian Institutes of Health Research to MRG.

This research was undertaken thanks also to funding from a Research Trainee Award from the Canadian Consortium on Neurodegeneration in Aging (CCNA) to NHMF.

