## Supplemental Table 1 for "Red flags for remote cognitive assessment: An expert consensus study using the Delphi method on behalf of the Canadian Consortium on Neurodegeneration in Aging"

Supplemental Table 1 – List of all the red flags with scores from the second round of the Delphi process. The flags are listed in the order they were shown to respondents. Data shown describes mean ± standard deviation.

| Red Flag | | QI1  Mean±SD | QI2  Mean±SD | QI3  Mean±SD | Overall  Mean±SD |
| --- | --- | --- | --- | --- | --- |
| 1 | The patient does not have access to a functional device that supports videoconferencing. | 5.0±0.0 | 4.8±0.4 | 3.9±1.5 | 4.5±0.9 |
| 2 | The patient is uncomfortable with using the technology. | 4.3±0.4 | 4.1±0.3 | 4.3±0.7 | 4.2±0.5 |
| 3 | The patient has significant anxiety from the computer screen or seeing themselves on the screen. | 4.4±0.7 | 3.6±0.7 | 3.4±1.1 | 3.8±0.9 |
| 4 | The patient has preference for in-person assessment. | 3.8±0.7 | 4.4±0.7 | 4.4±0.7 | 4.2±0.7 |
| 5 | The remote cognitive assessment is not in the best interest of the patient. | 4.5±0.7 | 3.1±1.2 | 3.4±1.0 | 3.7±1.0 |
| 6 | The patient does not have a long term residence or moves residences frequently. | 2.4±1.1 | 3.5±0.7 | 3.5±0.9 | 3.1±0.9 |
| 7 | The patient does not have sufficient ability to move around as needed (ex., the patient is bedbound.) | 2.3±1.0 | 3.4±1.1 | 3.6±1.2 | 3.1±1.1 |
| 8 | The patient has moderate-severe sensory impairment (ex., hearing loss). | 4.1±0.6 | 4.0±0.9 | 4.0±0.9 | 4.0±0.8 |
| 9 | The patient has severe anosognosia. | 2.8±1.4 | 3.1±0.8 | 2.5±0.5 | 2.8±1.0 |
| 10 | The patient has significant language dysfunction (ex., primary progressive aphasia) | 3.5±0.9 | 3.4±0.7 | 3.0±0.7 | 3.3±0.8 |
| 11 | The patient has severe visuospatial deficits. | 3.9±0.9 | 3.3±0.7 | 3.1±0.6 | 3.4±0.7 |
| 12 | The patient has severe behavioural issues. | 2.9±1.1 | 3.4±0.7 | 3.3±0.7 | 3.2±0.8 |
| 13 | The patient has mild-moderate, or worse, psychotic symptoms. | 2.9±1.2 | 3.8±0.7 | 3.3±0.7 | 3.3±0.9 |
| 14 | The patient has suicidal ideas. | 3.4±1.0 | 3.6±0.7 | 3.5±0.7 | 3.5±0.8 |
| 15 | The patient has a known medical or psychiatric condition that is decompensated. | 3.8±1.2 | 3.3±1.0 | 3.3±0.7 | 3.4±1.0 |
| 16 | The patient is suspected to have delirium or altered level of consciousness. | 4.6±0.5 | 3.4±1.4 | 3.4±1.3 | 3.8±1.1 |
| 17 | The patient has significant uncontrolled pain. | 3.3±1.3 | 3.5±1.1 | 3.8±1.0 | 3.5±1.1 |
| 18 | The patient is suspected to have a rapidly progressive neurodegenerative disease. | 3.1±1.3 | 3.4±1.0 | 3.3±1.2 | 3.3±1.2 |
| 19 | The patient has an atypical presentation that requires more thorough neuropsychological testing. | 3.8±0.8 | 3.6±0.9 | 4.5±0.5 | 4.0±0.7 |
| 20 | The patient is too cognitively impaired to understand the process of video assessment. | 3.9±0.6 | 3.0±1.0 | 3.0±1.2 | 3.3±1.0 |
| 21 | The patient is easily distracted, fidgeting, or walking away from the computer. | 3.3±0.8 | 2.8±1.2 | 4.1±0.6 | 3.4±0.9 |
| 22 | The patient consumes alcohol regularly or at unusual times of the day. | 3.4±0.9 | 3.6±0.7 | 3.8±1.2 | 3.6±0.9 |
| 23 | The patient has developed racial bias and is abusive towards the clinician. | 2.5±0.7 | 3.5±0.5 | 3.8±0.8 | 3.3±0.7 |
| 24 | There are physical manifestations accompanying the cognitive impairment, necessitating an in-person neurological exam. | 2.8±0.7 | 2.9±1.1 | 3.3±1.1 | 3.0±1.0 |
| 25 | There has been a sudden change in the patient's clinical status. | 4.0±0.7 | 3.1±1.1 | 3.3±1.1 | 3.5±1.0 |
| 26 | There is high caregiver burden or discomfort with using technology. | 4.1±1.3 | 4.3±0.4 | 4.4±0.5 | 4.3±0.8 |
| 27 | The caregiver has preference for in-person assessment. | 4.0±0.9 | 4.3±0.4 | 4.3±0.4 | 4.2±0.6 |
| 28 | The caregiver is opposed to cognitive assessment of the patient. | 3.3±1.1 | 3.5±0.9 | 3.5±0.9 | 3.4±0.9 |
| 29 | The patient does not have access to a caregiver during the remote assessment. | 3.5±1.1 | 4.1±1.1 | 4.4±0.7 | 4.0±1.0 |
| 30 | The patient does not have access to a caregiver during the remote assessment AND is more than mildly impaired. |  |  |  |  |
| 31 | The caregiver is unable to participate in a separate private interview. | 3.5±0.9 | 3.8±0.8 | 3.9±0.9 | 3.7±0.9 |
| 32 | The caregiver is a temporary caregiver. | 2.6±0.7 | 3.5±0.9 | 3.6±0.7 | 3.3±0.8 |
| 33 | The caregiver is suspected to have cognitive impairment. | 3.4±0.9 | 2.9±1.1 | 2.5±1.0 | 2.9±1.0 |
| 34 | The caregiver is interrupting the interview frequently. | 3.6±0.7 | 3.6±0.7 | 2.4±0.7 | 3.2±0.7 |
| 35 | There is suspicion of abuse (physical, psychological, sexual, financial) or neglect from the caregiver. | 4.8±0.4 | 3.3±0.8 | 3.0±1.1 | 3.7±0.8 |
| 36 | The caregiver is known to be unreliable. | 3.4±1.0 | 3.0±1.0 | 3.3±1.1 | 3.2±1.0 |
| 37 | The caregiver is known to have unstable medical or psychiatric disease. | 3.5±1.1 | 2.8±1.0 | 3.0±1.0 | 3.1±1.0 |
| 38 | The caregiver is showing signs of significant burnout. | 2.5±0.9 | 3.0±0.7 | 2.9±0.8 | 2.8±0.8 |
| 39 | The caregiver is uncomfortable discussing matters in front of the patient. | 3.3±1.1 | 3.5±0.5 | 3.5±0.9 | 3.4±0.9 |
| 40 | The caregiver has lots of concerns that need to be addressed. | 2.4±0.9 | 3.5±0.7 | 2.9±0.6 | 2.9±0.7 |
| 41 | The caregiver is distrustful of the clinician. | 3.4±0.9 | 3.0±0.7 | 2.9±0.8 | 3.1±0.8 |
| 42 | The caregiver is anxious. | 2.0±0.7 | 3.3±0.8 | 2.9±0.8 | 2.7±0.8 |
| 43 | The caregiver has racial bias towards the clinician. | 2.9±0.6 | 2.5±0.5 | 2.8±0.7 | 2.7±0.6 |
| 44 | The clinician does not have access to a device that supports videoconferencing. | 4.6±0.7 | 4.4±1.3 | 4.9±0.3 | 4.6±0.9 |
| 45 | The clinician has limited technical support for setup. | 4.0±1.0 | 3.9±0.9 | 4.4±0.5 | 4.1±0.8 |
| 46 | The clinician does not have experience with and knowledge of the benefits and limitations of remote assessment. | 4.1±1.1 | 3.9±0.9 | 4.1±0.6 | 4.0±0.9 |
| 47 | The clinician does not have access to the patient's prior chart or imaging. | 4.0±0.9 | 3.9±0.9 | 3.8±1.0 | 3.9±0.9 |
| 48 | The clinician does not have access to a service corridor for the patient. | 3.6±0.7 | 3.8±0.7 | 3.5±0.9 | 3.6±0.7 |
| 49 | The clinician feels they must censor themselves. | 3.4±1.1 | 3.3±0.7 | 3.1±0.6 | 3.3±0.8 |
| 50 | The clinician lacks experience with in-person cognitive assessment. | 3.9±1.3 | 3.8±1.0 | 3.8±1.0 | 3.8±1.1 |
| 51 | The clinician is distracted or lacks time to properly perform the assessment. | 4.1±0.9 | 3.6±1.1 | 4.0±1.0 | 3.9±1.0 |
| 52 | The clinician lacks expertise in neurocognitive disorders. | 4.3±1.0 | 3.9±1.5 | 4.5±0.5 | 4.2±1.0 |
| 53 | The clinician lacks the skills to diffuse unexpected behavior from the patient or caregiver. | 3.9±0.9 | 2.9±1.1 | 3.4±1.2 | 3.4±1.1 |
| 54 | The clinician lacks patience and empathy. | 3.5±1.0 | 2.8±1.1 | 3.1±1.1 | 3.1±1.0 |
| 55 | A private and safe spot for an assessment is not possible. | 4.9±0.3 | 4.4±1.0 | 4.1±1.1 | 4.5±0.9 |
| 56 | The patient or caregiver will not have technical support at the time of the encounter. | 3.4±0.9 | 3.9±1.1 | 3.6±1.1 | 3.6±1.0 |
| 57 | The patient does not have access to a quiet space without distractions. | 5.0±0.0 | 4.4±0.5 | 4.0±0.9 | 4.5±0.6 |
| 58 | The patient's environment contains orienting cues. | 3.3±0.7 | 3.5±0.9 | 3.1±0.8 | 3.3±0.8 |
| 59 | There are language barriers, or the standardized assessments are not available in the patient's native language. | 3.9±0.6 | 4.1±0.3 | 4.0±0.0 | 4.0±0.4 |
| 60 | It is the first diagnostic assessment to address cognitive or behavioural concerns. | 2.5±0.9 | 4.4±0.5 | 4.1±0.9 | 3.7±0.8 |
| 61 | The patient scores close to cutoff on a standardized cognitive assessment. | 2.8±1.0 | 3.9±0.6 | 3.9±0.8 | 3.5±0.8 |
| 62 | The patient is being seen for a second opinion. | 2.6±1.1 | 4.1±0.6 | 4.1±0.6 | 3.6±0.8 |
| 63 | The patient is being seen for medico-legal reasons (ex., court order) | 4.0±0.7 | 4.3±0.7 | 4.3±0.7 | 4.2±0.7 |
| 64 | The assessment is at a time of day when the patient might not be awake or alert. | 3.3±1.1 | 4.0±1.0 | 3.6±1.1 | 3.6±1.1 |
| 65 | Disclosure of the diagnosis or consequences thereof is needed during the encounter. | 2.5±0.7 | 3.9±0.3 | 3.4±0.9 | 3.3±0.7 |
| 66 | The patient is outside the legal jurisdictions of the clinician's medical license. | 5.0±0.0 | 4.5±1.0 | 4.9±0.3 | 4.8±0.6 |
| 67 | It would be impossible to eventually examine the patient in-person. | 2.9±1.3 | 4.1±0.8 | 3.9±1.1 | 3.6±1.1 |
